# Genetic risk factors and Covid-19 severity in Brazil: results from BRACOVID Study

**DOI:** 10.1101/2021.10.06.21264631

**Authors:** Alexandre C. Pereira, Taniela Marli Bes, Mariliza Velho, Emanuelle Marques, Cintia E Jannes, Karina Ramos Valino, Carla L Dinardo, Silvia Figueiredo Costa, Alberto José da Silva Duarte, Alexandre Ricardo dos Santos, Miguel Mitne-Neto, Jose Medina-Pestana, Jose E Krieger

**Author notes:** Corresponding author. (ACP), (JEK).

## Abstract

The Covid-19 pandemic has changed the paradigms for disease surveillance and rapid deployment of scientific-based evidence for understanding disease biology, susceptibility, and treatment. We have organized a large-scale genome-wide association study in Sars-Cov-2 infected individuals in Sao Paulo, Brazil, one of the most affected areas of the pandemic in the country, itself one of the most affected in the world. Here we present the results of the initial analysis in the first 5,233 participants of the BRACOVID study.

We have conducted a GWAS for Covid-19 hospitalization enrolling 3533 cases (hospitalized Covid-19 participants) and 1700 controls (non-hospitalized Covid-19 participants). Models were adjusted by age, sex and the 4 first principal components. A meta-analysis was also conducted merging BRACOVID hospitalization data with the Human Genetic Initiative (HGI) Consortia results.

BRACOVID results validated most loci previously identified in the HGI meta-analysis. In addition, no significant heterogeneity according to ancestral group within the Brazilian population was observed for the two most important Covid-19 severity associated loci: 3p21.31 and Chr21 near IFNAR2. Using only data provided by BRACOVID a new genome-wide significant locus was identified on Chr1 near the genes DSTYK and RBBP5. The associated haplotype has also been previously associated with a number of blood cell related traits and might play a role in modulating the immune response in Covid-19 cases.

## Introduction

The severe acute respiratory syndrome coronavirus 2 (SARS-CoV-2) was first described in Wuhan, China, in December 2019, and quickly spread across the globe causing a pandemic (1). After 20 months of the beginning of the spread, there were more than 213 million confirmed cases worldwide, with total deaths exceeding 4 million, half million in Brazil alone (2).

Clinical outcomes of SARS-CoV-2 infection range from asymptomatic infection to fatal coronavirus disease 2019 (Covid-19). The high mortality rates are predominantly among specific subgroups with predeterminate conditions, such as male sex, older age or having pre-existent cardiovascular, pulmonary, or immune disease. However, many severe clinical cases are not due to comorbidity or constitutional factors. Notably, even among those without pre-existing conditions, inter-individual heterogeneity is striking. The considerable variation in disease behavior among infected individuals highlights the importance of the hosts’ genetic background modulating disease presentation (3–5).

Until now a small number of large-scale genome-wide association studies (GWAS) has been conducted with some reproducible identified genetic loci associated with disease susceptibility or severity. A GWAS among 1980 patients from the European Covid-19 epicenters in Italy and Spain, as well as from the less-burdened countries of Germany and Norway was the first study to describe the susceptibility locus at the chromosome 3p21.31 gene cluster and suggested the involvement of the ABO blood-group system in Covid-19 (6). The *GenOMICC* study, in addition to replicating the association at chromosome 3p21.31 as mediator of critical illness, identified associations with disease severity and other genetic variants (7). Finally, early in 2021, the Covid-19 Host Genetics Initiative (HGI) has brought together 49.562 cases and 2 million controls, from populations with different ancestries across the world to elucidate the genetic determinants of SARS-CoV-2 infection and severity including 853 samples from the BRACOVID study. From these analyses 15 genome-wide significant loci associated with SARS-CoV-2 infection or Covid-19 were observed (8).

Despite these efforts, the still limited number of identified loci that appears to modulate the relationship between clinical severity and human genetic variation precludes a thoroughly understanding of inter-individual susceptibility in the disease. In addition, most studies are overwhelmingly represented by European or Asian individuals and whether the identified loci are also operant on individuals from other ancestry backgrounds is unknown. Here we present the results from the BRACOVID study enrolling 5,233 participants with distinct COVID-19 clinical presentations and from a diverse population in Brazil. Our results confirm most previously identified loci and add a new genome-wide significant locus associated with COVID-19 hospitalization in the Brazilian population.

## Results

The BRACOVID study was designed to identify and characterize genetic risk factors for Covid-19 severity in the Brazilian population.

### Relationship between demographic and clinical factors with Covid-19 hospitalization

We summarize the demographic, clinical and laboratory characteristics of the 5,233 participants of the study in Table 1 according to hospitalization necessity.

**Table 1.**
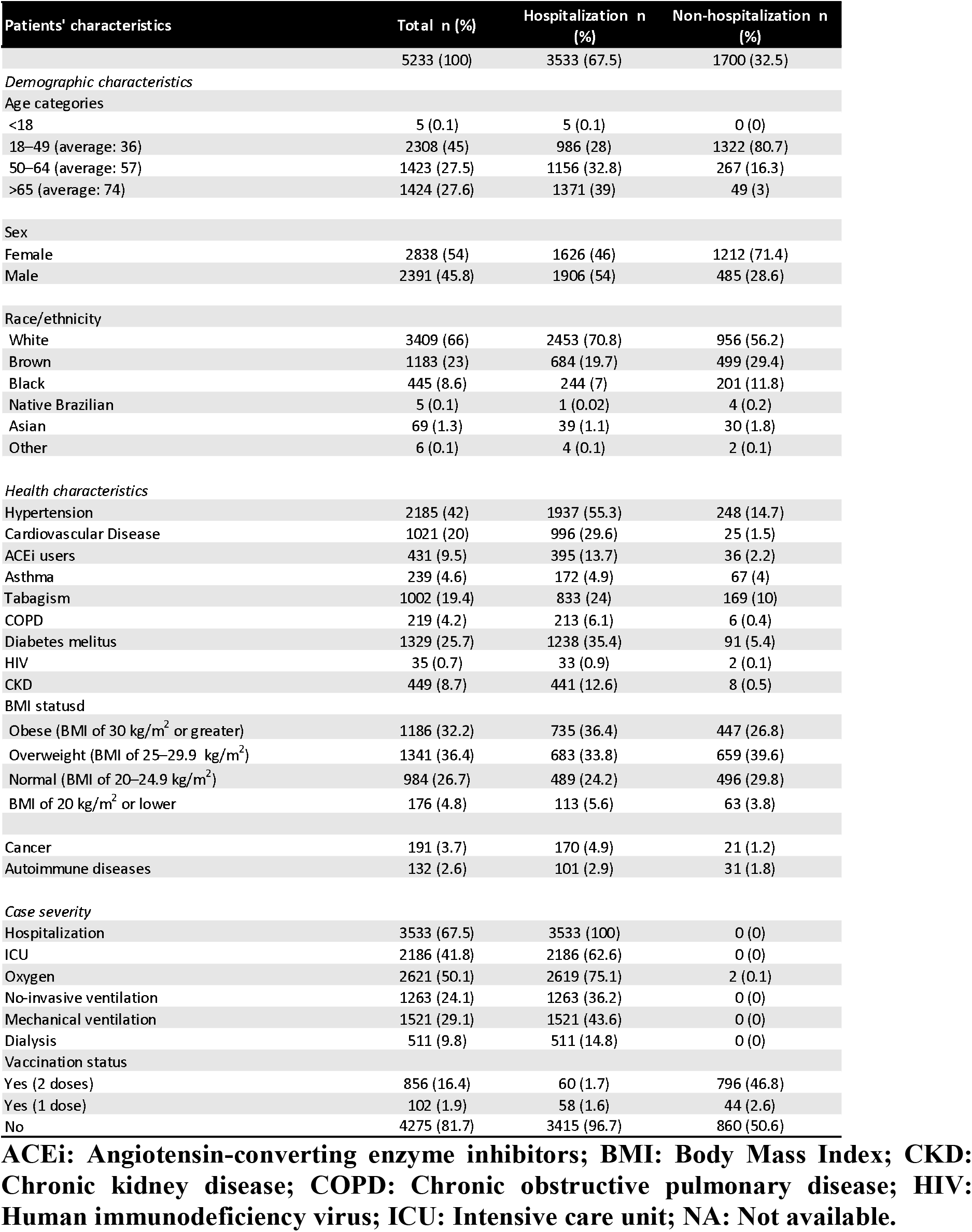
Demographic, clinical and laboratory characteristics of studied subjects according to hospitalization need.

### Genome-Wide Association Analysis of Covid-19 hospitalization

We have performed a GWAS of the genetic association architecture regarding Covid-19 hospitalization need. Groups were 1,700 individuals that did not require hospital admission due to Covid-19 and 3,533 individuals hospitalized due to severe Covid-19. In the primary analysis, we have run 2 GWAS analysis (one for each used genotyping array). Each analysis was adjusted for the first 4 Principal Components, age, and sex. GWAS results were then merged using the meta-analysis routine in plink. The observed genomic inflation factor for the adjusted analysis was lambda = 1.05.

In our main analysis, we observed one genome-wide significant locus that reached the pre-defined genome-wide significant level of 5 × 10^−8^ (Figure 1). We also observed four additional loci that reached the pre-defined p-value threshold of 1 × 10^−6^ (Table 2 and Supplementary Figure 1). We have run sensitivity analysis stratifying individuals by their main genetic ancestral component and then conducting a trans-ethnic meta-analysis. Results were very similar to those observed in the main analysis (Supplementary Figures 2 and 3).

**Table 2.** Genome-wide significant and suggestive loci identified in BRACOVID Covid-19 severity analysis.

| CHR | BP | SNP | Effect Allele | P | OR | Genes |
| --- | --- | --- | --- | --- | --- | --- |
| 1 | 205208489 | chr1:205208489:G:A | G | 3.99E-08 | 1.3516 | RBBP5, DSTYK, TMCC2 |
| 2 | 45584979 | chr2:45584979:T:C | C | 4.06E-07 | 0.7509 | SRBD1 |
| 4 | 25613313 | chr4:25613313:G:A | A | 3.21E-07 | 1.4647 | SLC34A2 |
| 5 | 21768177 | chr5:21768177:G:A | A | 5.02E-07 | 1.3263 | GUSBP1, CDH12 |
| 10 | 22439346 | chr10:22439346:T:C | C | 5.96E-07 | 0.549 | PIP4K2A |

**Figure 1.**
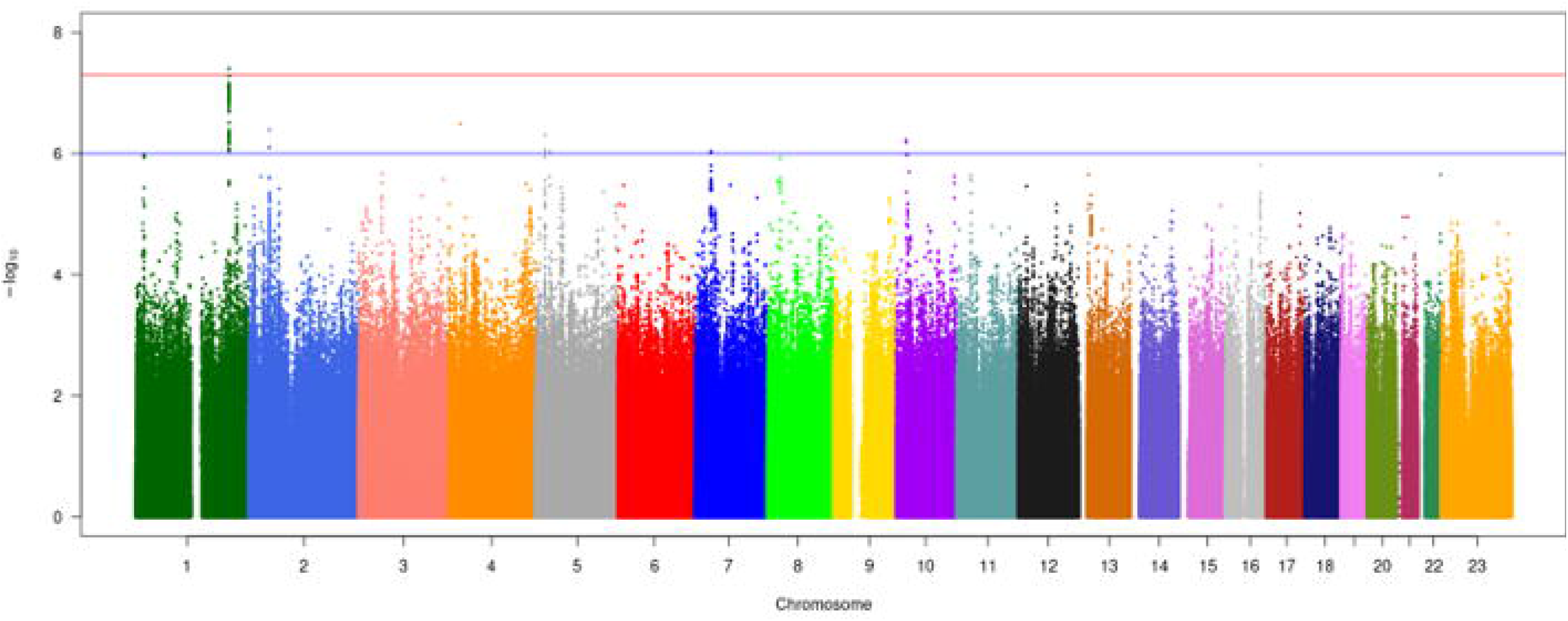
Manhattan plot of BRACOVID results for Covid-19 hospitalization. Results from a logistic regression model adjusted for the first 4 PCs, age, and sex for each used genotyping array were meta-analyzed using a fixed-effect meta-analysis.

### BRACOVID HGI meta-analysis

We have replicated most previously described HGI (8) (B1 analysis) results (BRACOVID was not part of this specific HGI sub-analysis) (Supplementary File 2). Specifically, both loci at 3p21.31 and Chr21 near IFNAR2 previously associated with hospitalization need on HGI B1 analysis were nominally significant in our study with the same directionality of effect. We have also compared our results to those described by HGI B2 analysis. We have observed significant associations in all previously described genome-wide loci but for the one on chr9, near the ABO locus (Supplementary File 3).

In addition to looking for replicated signals within our results, we ran two meta-analyses between BRACOVID hospitalization need and HGI B1 and B2 analysis. Adding our results to those provided by HGI B1 analysis disclosed a genome-wide association signal on chr6 that was only previously seen on the HGI B2 analysis, strengthening the confidence that this locus is indeed associated with Covid-19 severity (Supplementary File 4 and Supplementary Figure 4).

Our results meta-analyzing BRACOVID summary statistics with HGI B2 results supported most previously described genome-wide association loci. Namely, for Chr1 near THBS3; Chr3 LZTFL1-CXCR6; Chr6 HLA-DRB1; Chr6 FOXP4; Chr11 MUC5B; Chr11 ELF5; Chr12 OAS2; Chr12 P2RX2; Chr17 CRHR1; Chr19 POLD1; Chr19 TYK2; and Chr21 IFNAR2 regions we were able to observe concordant signals and no heterogeneity of effects. We want to emphasize the observation of significant heterogeneity for the previously described loci on Chr9 ABO locus; Chr21 SLC5A3; and Chr17 TAC4 suggesting that these may indeed be false-positive associations due to population structure confounding, population-specific loci or loci associated with Covid-19 susceptibility and not severity (Supplementary File 5).

### Local association structure suggests a population specific effect for Chr1 DSTYK locus

The newly identified genome-wide significant locus near DSTYK was not observed as significantly associated in our HGI meta-analysis. By comparing the local association structure at the DSTYK locus, it is interesting to note that HGI B2 results also suggest a sub-genome-wide significant association at the same region (Figure 2A and B). However, albeit in the same recombination interval, there is no significant colocalization of the two genetic association structures, suggesting (1) the existence of more than one causal variants within this region or (2) that different LD structures among human populations may be confounding the association pattern at this locus.

**Figure 2.**
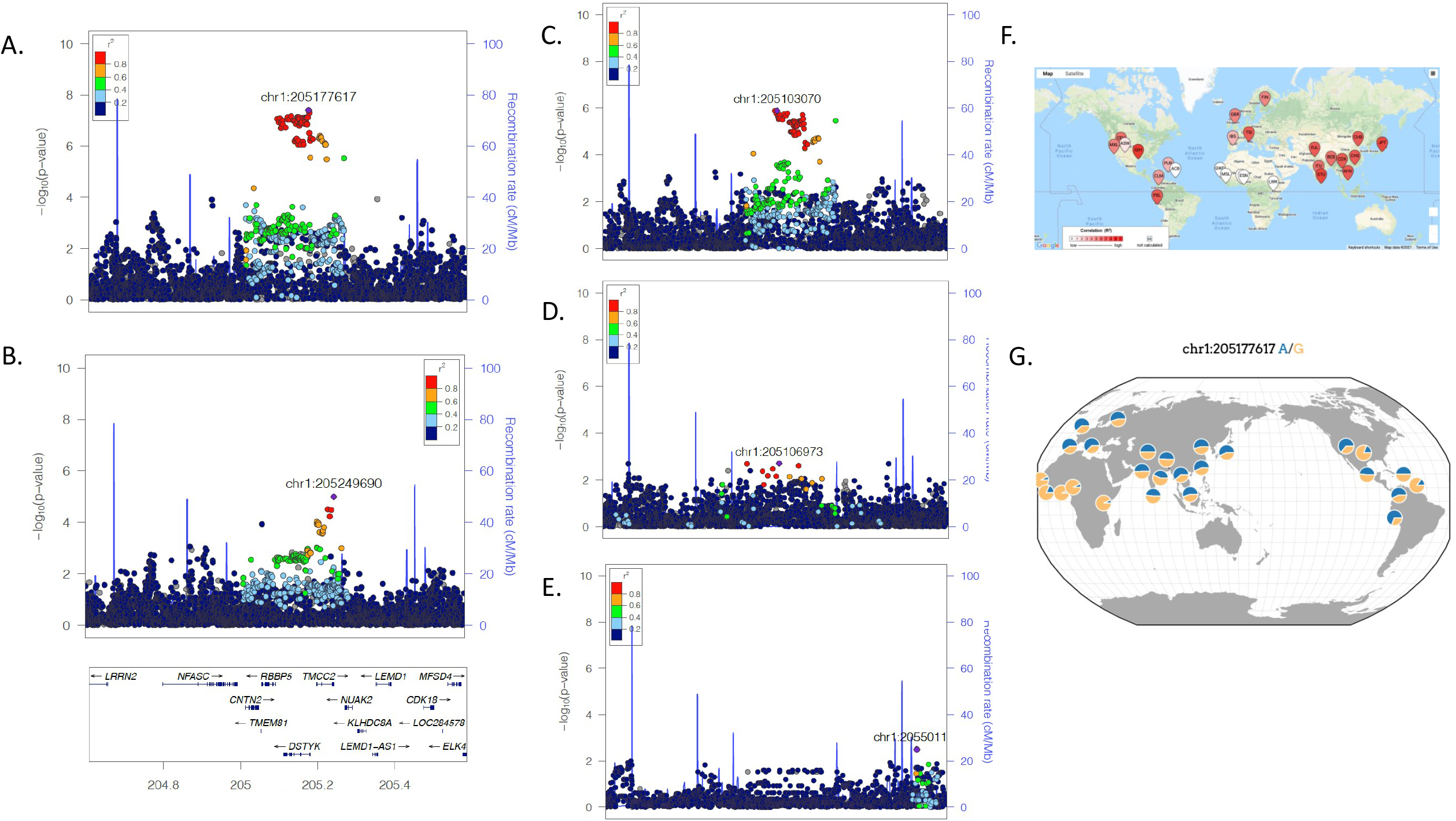
Association and LD structure at Chr1 DSTYK associated locus. A. Local association structure of BRACOVID hospitalization analysis; B. Local association structure of HGI B2 analysis; C. Local association structure for BRACOVID European cluster; D. Local association structure for BRACOVID African cluster; E. Local association structure for BRACOVID Native American cluster; F. World-wide distribution of pairwise LD between rs11240388 (local maximum for BRACOVID analysis) and rs9661015 (local maximum for HGI B2 analysis); G. 1000G allele frequency distribution for rs11240388.

To explore these scenarios, we have first analyzed the local association structure within the Brazilian sample stratifying by different degrees of European, African, and Native American genetic ancestry among participants. Curiously, although the signal is shared among the 3 groups, the strongest association signal was observed among the Brazilian individuals with high European genetic ancestry (Figure 2C). In addition to this, we have studied the pairwise LD between the most associated SNV in the Brazilian analysis (rs11240388), and the most associated SNV in HGI B2 analysis (rs9661015). Strikingly different world-wide pairwise LD patterns are observed among these 2 markers (Figure 2F, https://ldlink.nci.nih.gov/). In our sample, the R-square between the two markers was 0.32 for participants with high European ancestry, and 0.11 for participants with high African ancestry.

Using only individuals with high European ancestry from our Brazilian dataset we delineated the associated haplotype boundaries between chr1:205131352 and chr1:205210329, defining a haplotype of approximately 79Kb. Looking into the haplotype structure of markers part of the associated haplotype we did not observe a significant difference in their pairwise LD structure among Brazilians with high European or high African ancestry. Using rs11240388 as a tag-SNP for our risk haplotype, we defined world-wide allele frequencies for the risk haplotype (Figure 2G, https://popgen.uchicago.edu/ggv/). In our own sample, the frequency of the G allele (risk allele) was 48% for the entire sample, 42% in Brazilian individuals with high European ancestry and 59% in Brazilian individuals with high African ancestry. Despite having increased frequency in Brazilian individuals with high African ancestry, and as expected from the stratified local association plots, the haplotype was only significantly associated with Covid-19 severity in Brazilian individuals with high European genetic ancestry. Taken together these results suggest that the causal variant responsible for the association is present in a 79Kb haplotype derived from European populations.

### rs11240388 is also associated with several blood cell traits and with DSTYK and RBBP5 gene expression levels

Exploring the genomic annotations for the newly observed locus associated with Covid-19 hospitalization we used rs11240388 as a proxy of the risk haplotype for it is in complete LD with all the markers of the 79Kb haplotype. Rs11240388 is located at chr1:205208489 within the DSTYK gene region. The region appears to be delimited by a recombination region spanning the genes CNTN2, TMEM81, RBBP5, DSTYK, and TMCC2 and harbors several GeneHancer regulatory elements, most notably for CNTN2, DSTYK and TMCC2 (Figure 3A). Querying the MR-base database for other phenotypes previously shown to be associated with rs11240388, we observed 69 traits (Figure 3B). Of particular interest to Covid-19 biology are blood cell related traits and lung associated traits (Supplementary File 6).

**Figure 3.**
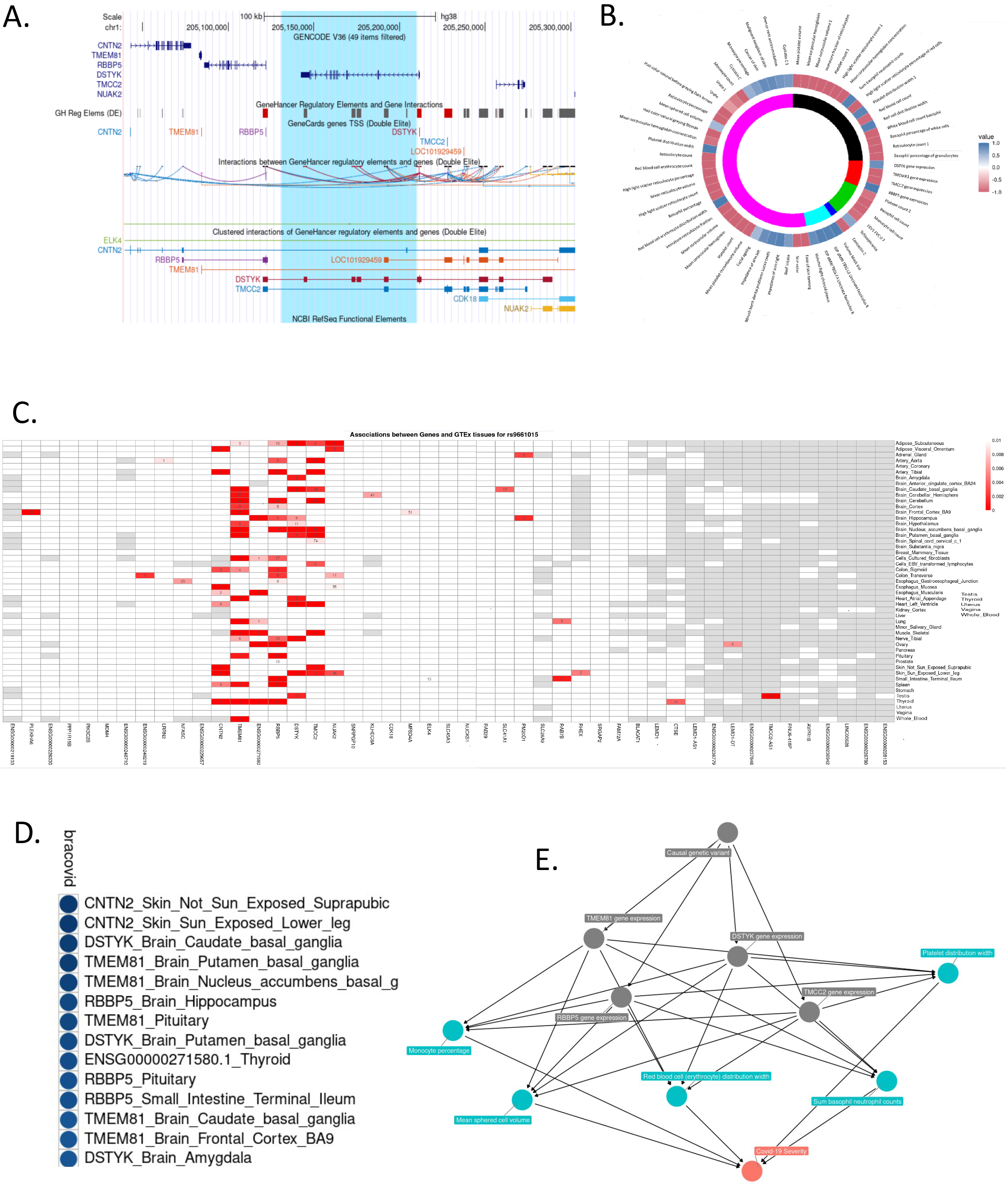
Annotation and functional characterization of Chr1 DSTYK associated region. A. Genomic anatomy of associated region. Blue highlight – 79Kb associated haplotype. B. Phewas results and colocalization analysis with BRACOVID summary statistics. Colocalization values are depicted as a function of their H3 and H4 posterior probabilities. Positive values (blue spectrum) for higher H4 values than H3, negative values (red spectrum) for higher H3 than H4. Inner circle represents study/database of trait being colocalized with BRACOVID results. Black: Datasets that satisfy minimum requirements imported from the EBI database of complete GWAS summary data; red: eqtlGen 2019 results; green: GWAS summary datasets generated by many different consortia that have been manually collected and curated, initially developed for MR-Base (round 2); dark blue: Complete GWAS summary data on protein levels as described by Sun et al 2018; magenta: Complete GWAS summary data on brain region volumes as described by Elliott et al 2018; pink: Neale lab analysis of UK Biobank phenotypes, round 2; C. Association structure between rs11240388 and genes within its genomic region using 42 tissues in GTEx V8. Red rectangles are for association p values below 0.01. Numbers within cells represent the normalized mean expression value within tissue for gene; D. Colocalization results between BRACOVID summary statistics for rs11240388 region and different GTEx V8 gene/tissue combinations. Shown are only the most significant H4 probability results; E. Causal diagram derived from pairwise comparisons with significant colocalization at rs11240388 region.

Next, we explored the association structure between rs11240388 and GTEx gene expression levels in all tissues and for all genes within the region (Figure 3C). Results from GTEx suggest that rs11240388 is an eQTL in several tissues for CNTN2, TMEM81, RBBP5 and DSTYK. Colocalization analysis between GTEx association results and BRACOVID association results confirmed a significant colocalization for these genes in several different tissues, suggesting that the association between the 79Kb risk haplotype and Covid-19 severity might be mediated through gene expression modulation in the genes located in this genomic region (Figure 3D). Finally, we tested all pair-wise possibilities for colocalization: BRACOVID signal, the 69 associated traits for the same region, and all GTEx gene-tissue results with significant colocalization (higher than 0.7) with BRACOVID (Supplementary Figure 5). This analysis provides several potential intermediate mediators that could explain the association between the identified risk haplotype and Covid-19 severity (Figure 3E).

## Discussion

Covid-19 has been a world-wide pandemic for more than one year. The disease has killed more than 4 million individuals and has de-stabilized entire health systems as well as the world economy (2,9,10). One striking characteristic of the disease is the large inter-individual variability regarding its severity, ranging from asymptomatic to fatal. Although several identified factors influence disease severity, predicting the overall Covid-19 outcome in a particular individual remains a challenge (3–5).

Since the beginning of the pandemic, several groups worldwide have joined efforts to uncover host factors. The Host Genetic Initiative (HGI) included researchers and samples from different parts of the globe to focus on the inter-individual variability in disease presentation (8) and outcomes and the BRACOVID is currently the largest such effort in Latin America.

BRACOVID was designed to compare individuals with mild or asymptomatic Covid-19 with those with severe Covid-19 requiring hospitalization. This contrasts with most genetic studies on host determinants of Covid-19, that use individuals from the general population as their main control group. As such, BRACOVID has a more specific study protocol aimed at identifying host genetic determinants of disease severity, and not susceptibility.

The results here presented replicate most, but not all, previously reported genome-wide significant loci associated with Covid-19 severity. These came from 3 prior publications: The Severe Covid-19 GWAS Group, *GenOMICC* and HGI (6–8). It is interesting to note that despite the large sample sizes, none of these prior studies have in fact compared individuals with mild versus severe Covid-19. As such, our results confirm some of the initial results regarding Covid-19 genetic determinants in a more diverse population and, most importantly, within a study design to differentiate between markers of susceptibility to infection and predictors of severity. Also of importance, although BRACOVID has participated in the meta-analysis of both *GenOMICC* and HGI, due to the analytical plan of these two prior publications, the contrast used by BRACOVID was not the one presented here. Specifically, in both *GenOMICC* and HGI, BRACOVID summary statistics were obtained by comparing hospitalized Covid-19 individuals with individuals from the general Brazilian population (the last group, not included in the present analysis) on what is called the HGI B2 analysis. Even in this latter situation, BRACOVID sample of hospitalized individuals used in the last version of HGI analysis, the one used in our meta-analysis with HGI B2, was of only 853 individuals, in contrast to the more than 3,500 hospitalized participants used in the present report.

Besides the importance of showing replications to previously identified loci in an admixed population, understanding loci that do not replicate might also be important in the follow-up efforts to identify population specific determinants of Covid-19 severity. One of the loci that was not replicated in our analysis is the association signal initially identified in the ABO locus. Our results, together with the verification of intense heterogeneity in signal described by HGI, suggest that the association initially described at this locus may be due to confounding by population structure or other higher order effects.

We have observed a new genome-wide significant locus associated with Covid-19 in our population. It is comprised by a 79Kb haplotype located on chromosome 1 near the DSTYK gene. The association signal in our sample was only observed in individuals with high European ancestry and was not described by previous studies, suggesting it might harbor a population specific risk allele or haplotype. Indeed, leveraging data from the 1000G project (11), we observed great inter-population differences in the haplotype frequency (although an apparently constant LD structure in the region among human populations).

Leveraging data from GTEx we were able to associate this haplotype with gene expression levels of at least 5 genes: DSTYK, TMEM81, RBBP5, CNTN2 and TMCC2. In addition, exploring prior publicly available GWAS data we found significant associations between the identified risk haplotype and 69 different traits (8). From these, using colocalization analysis, we have observed significant colocalization between BRACOVID results and several of these 69 traits, most strikingly a number of hematological traits, including immune cell morphology and number.

The main limitation of this study is the lack of a suitable replication sample for the GWAS results. The observed genome-wide significant locus near DSTYK needs to be replicated in an independent sample to continue to be considered a modulator of Covid-19 severity. The continuous effort to increase the sample size of the BRACOVID study may allow the identification genome-wide significant loci with decreased effect size.

Taken together, our analysis supports the hypothesis that the identified 79Kb haplotype on chromosome 1 may modulate Covid-19 relevant immunological traits through the gene expression of at least one of the genes with significant eQTLs in the region.

## Materials and methods

### Study population

The study sample belongs to the BRACOVID Study. For the present analysis we used the first 5,233 participants enrolled in the study.

The BRACOVID study was designed to identify and characterize genetic risk factors for Covid-19 severity in the Brazilian population. Participants were selected through two different ascertainment processes. First, Covid-19 patients were enrolled after hospitalization in one of the following tertiary care centers in the metropolitan area of Sao Paulo, Brazil: Instituto do Coração, and Instituto Central do Hospital das Clínicas da Faculdade de Medicina da Universidade de São Paulo. Non-hospitalized cases were selected by serological studies surveys for previous SARS-CoV-2 infection or SARS-CoV-2 PCR test among health professionals or the general population.

After signing an informed consent, a sample of whole-blood already collected for in-hospital biochemical analysis or SARS-CoV-2 serology was used for genomic DNA extraction.

Participants responded to a detailed questionnaire aimed at collecting demographic, risk factors, symptoms, and prior clinical information. For hospitalized participants we extracted the data from the hospital EHR.

### SNP Genotyping and Imputation

Genomic DNA extraction has been previously described (12). BRACOVID DNA samples were genotyped using two different Axiom arrays: Axiom_PMRA.r3 array (N=2605) or the Axiom_sarscov array (N=2628) (ThermoFisher, Waltham, USA) and genotypes annotated using the array specific annotation file provided at the ThermoFisher website. Genotype calling was performed using Affymetrix Power Tools. Initial VCF file contained 850483 (for the PMRA array) and 779972 (for the Sars-Cov array) variants before quality control filtering.

Imputation was performed using the Haplotype Reference Consortium Michigan Imputation Server using the TOPMED reference haplotype panel as reference (for mixed samples). More specifically, the Michigan Imputation Server used Minimac4 to conduct imputation on 658,357 SNPs remaining after data quality control. After imputation data were exported in the standard PLINK format, downstream QC procedures and statistical analysis were conducted using the latest PLINK (http://pngu.mgh.harvard.edu/_purcell/plink) and R software packages (http://www.r-project.org/), installed on a Linux based computation resource. Specifically, imputation markers were kept if R2 > 0.3, and minor allele frequency (MAF) > 0.01. A HWE p-value <1 × 10^−20^ was used to control for potential genotyping clustering problems. Genetic population structure was determined through PCA analysis after LD-pruning of associated markers (see also Statistical Analysis section). A total of 11,395,074 SNPs were used for genome-wide analyses, 10,981,197 for autosomal, and 413,877 for X-chromosomal analysis.

### GWAS Analysis

We used a dichotomous category defined among SARS-CoV-2 participants. We have grouped individuals that did not require hospitalization into the control group (participants ascertained through population-based serological prevalence studies that did not have a prior history of hospitalization due to Covid-19) and hospitalized individuals (ascertained at the first hospital admission day) in the case group. Baseline categorical parameters are presented using frequencies (proportions), continuous parameters are presented using mean ± SD.

Genome-wide association analyses were conducted using plink. We have conducted two analyses one without any further adjustment and one adjusting for the first four principal components, age, sex, and array type. The threshold for genome-wide significance was set to *p* <5×10^−8^. Associations with *p* <1×10^−6^ were considered as suggestive and presented as a list of top associated SNPs.

Due to the high level of admixture and complex genetic population structure present in the Brazilian population we have conducted different sensitivity analysis taking into consideration the individual position on the PCA plot generated using the 2 first principal components. Briefly, for the PCA-defined subgroup analysis we have k-means clustering with k=3 and defined three different subgroups with higher European, African, and Native-American ancestries. Meta-analysis used a fixed-effect model and was calculated using plink –meta-analysis routine.

Local association plots were created using LocusZoom (13). Local linkage disequilibrium structure was determined using Haploview (14).

### PheWas analysis

After identifying SNV that could be proxies of the associated haplotype we have used MR-base through the R package *ieugwasr* to search for prior GWAS studies reporting significant levels of association between a trait and the selected tag-SNV. The used p value cut-off level for this phewas analysis was *p* <1×10^−5^.

### Colocalization analysis

For colocalization analysis we have defined a window spanning 400Kb centered at the most associated variant in all regions classified as having a suggestive association signal. Information on all variants within this region was used for colocalization testing. We have used the R package *coloc* for colocalization analysis. Briefly, all genes residing in each selected region with their expression quantitative trait loci (eQTL) summary statistics available in GTEx V8.0 were sequentially tested for colocalization with the results obtained for Covid-19 severity association. As reference LD structure we used 1000 genomes 2012 European LD matrix (our sample has approximately 80% European ancestry). Colocalization was tested against all 48 tissues available in GTEx V8.0 and against all traits with significant association to the selected variant (as described in the phewas section). We used a threshold of H4 (the posterior probability that a single causal variant, or haplotype, could explain the local association pattern of both tested traits) > 0.7 as evidence for significant colocalization.

## Data Availability

The data regarding this study are available with the manuscript file.

## Acknowledgements

The BRACOVID project received an unrestricted financial support from JBS S.A. Company, Sao Paulo, Brazil and from the general public under the HC-COMVIDA crowdfunding scheme (https://viralcure.org/c/hc). The funds were managed by Fundação Zerbini and Fundação Faculdade de Medicina, respectively. We are thankful for the infrastructure support for sample collection provided by the HCFMUSP Covid-19 Study Group. We are also grateful to the Host Genetic Initiative for making their data publicly available (full acknowledgements can be found at https://www.covid19hg.org/acknowledgements/).

## Supporting Information

### Supplementary Figures

**Supplementary Figure 1.**
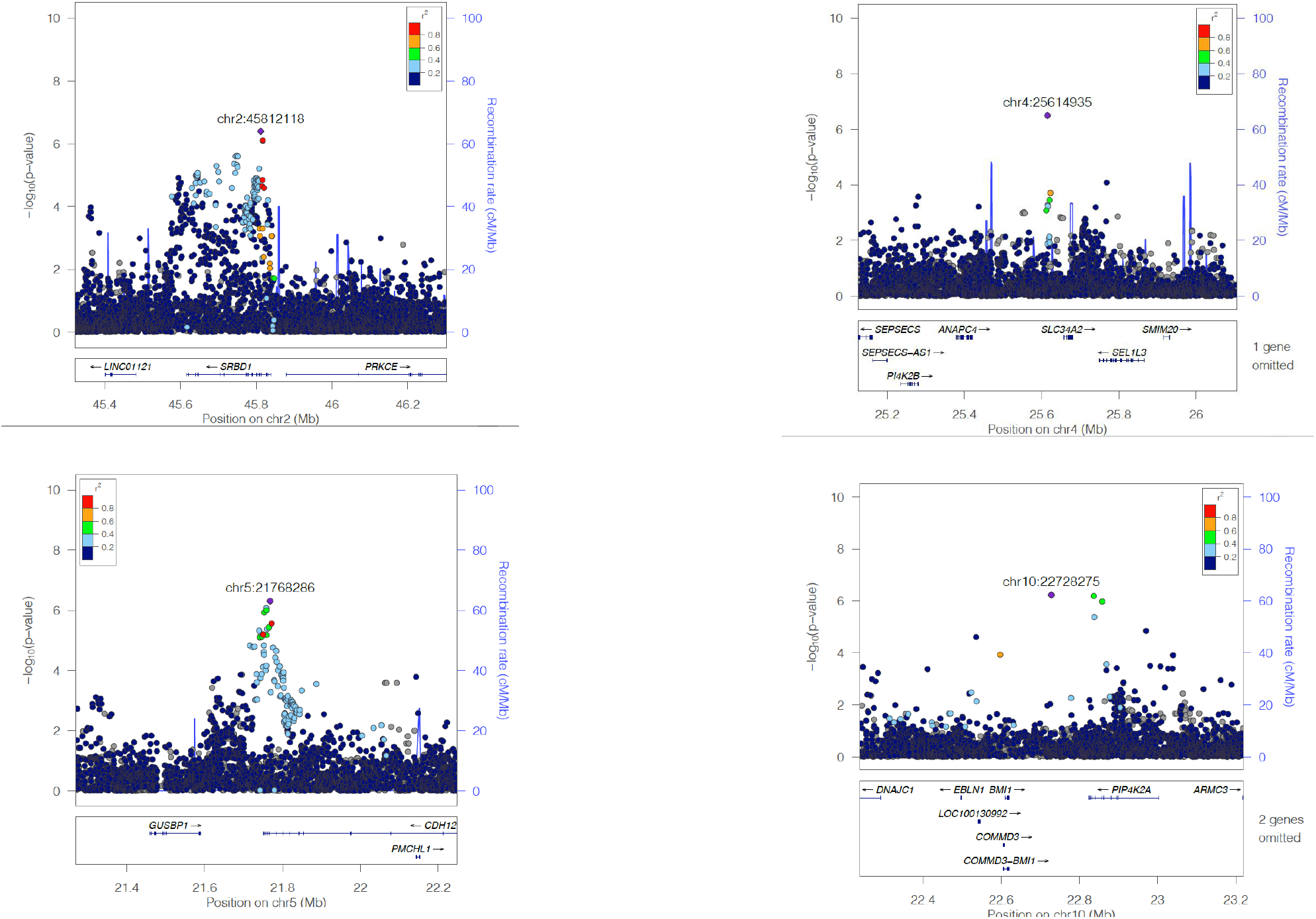
Local association plots for the 4 suggestive associated loci. Suggestive association was defined as a p-value between 1 × 10-6 and 5 × 10-8. Genome positions in Hg19.

**Supplementary Figure 2.**
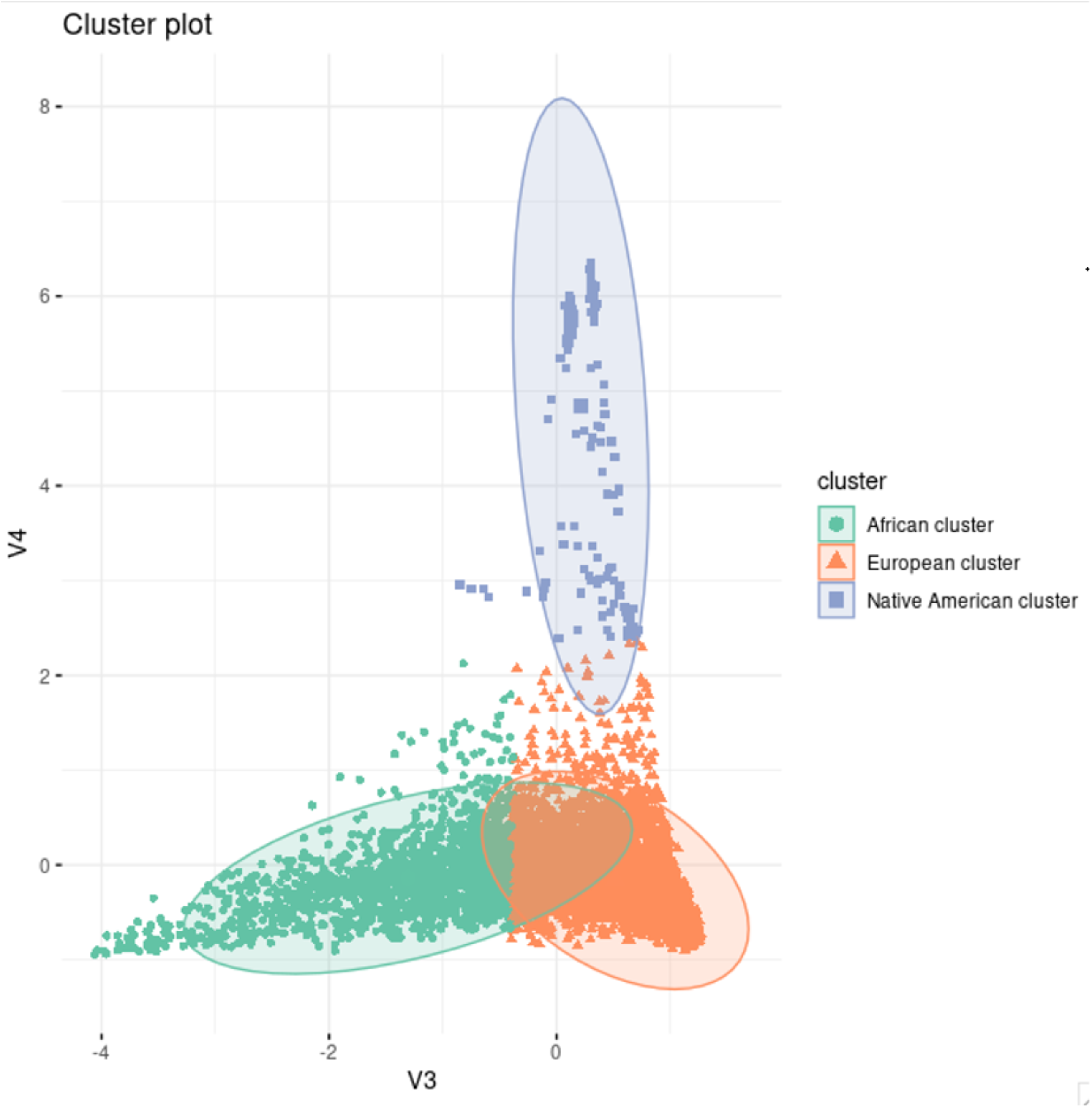
K-means clustering (k=3) of BRACOVID participants using the first and second PCs. Orange triangle - European cluster; Green circle – African cluster; Purple square – Native American cluster.

**Supplementary Figure 3.**
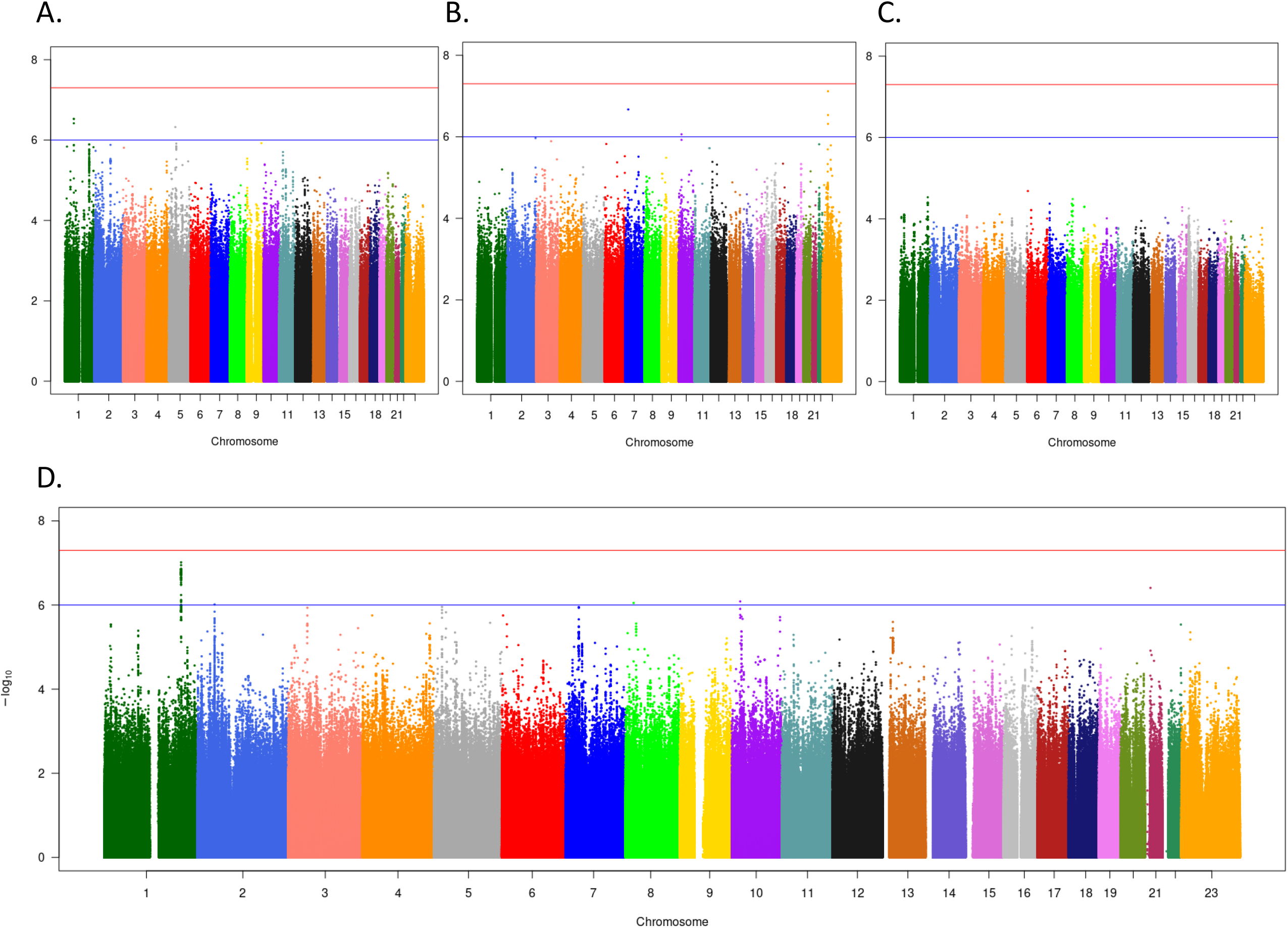
Manhattan plots of ancestry clusters and trans-ethnic meta-analysis. A. European cluster; B. African cluster; C. Native American cluster; D. Native American cluster.

**Supplementary Figure 4.**
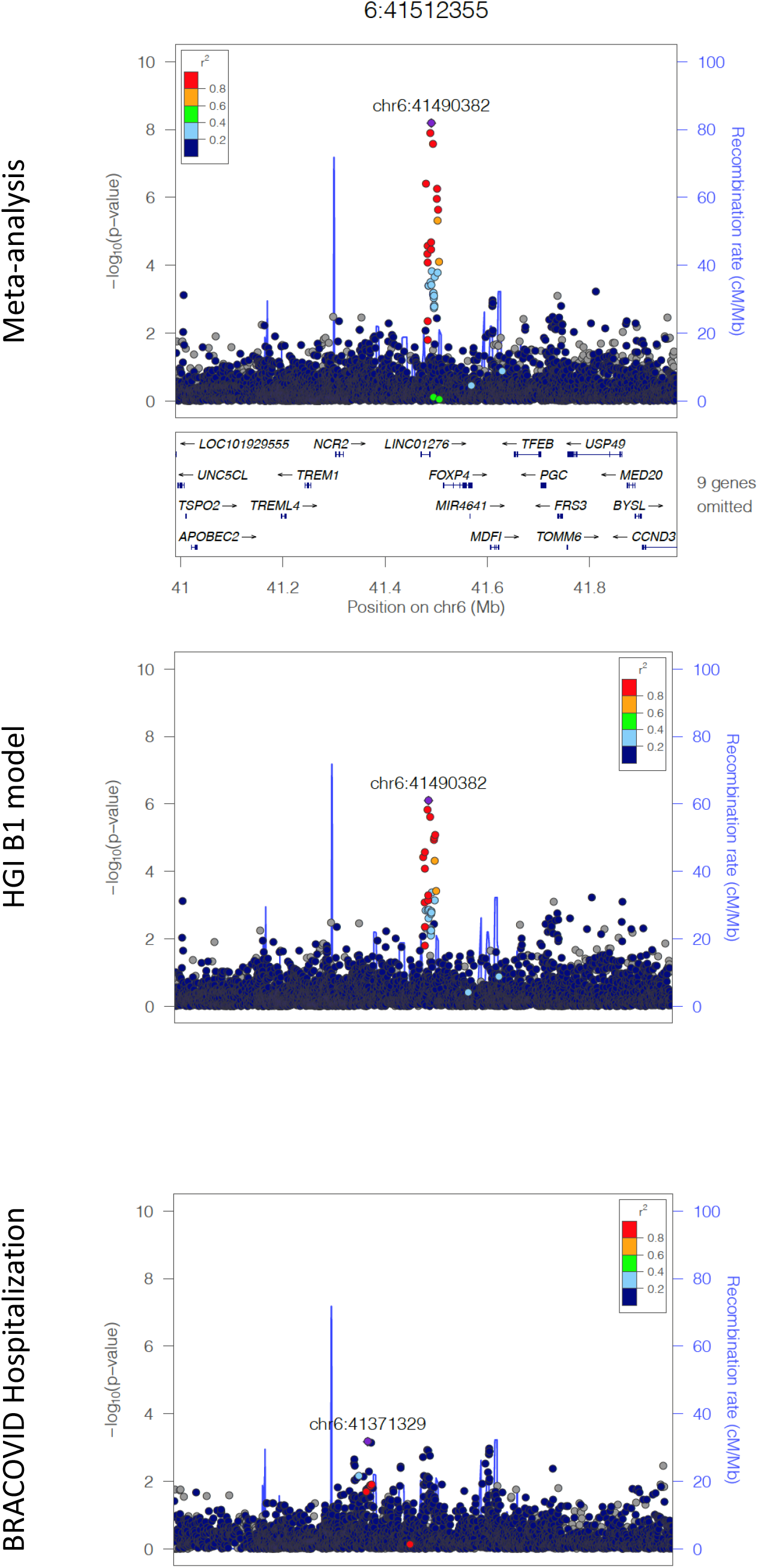
Local association structure for Chr6 FOXP4 region.

**Supplementary Figure 5.**
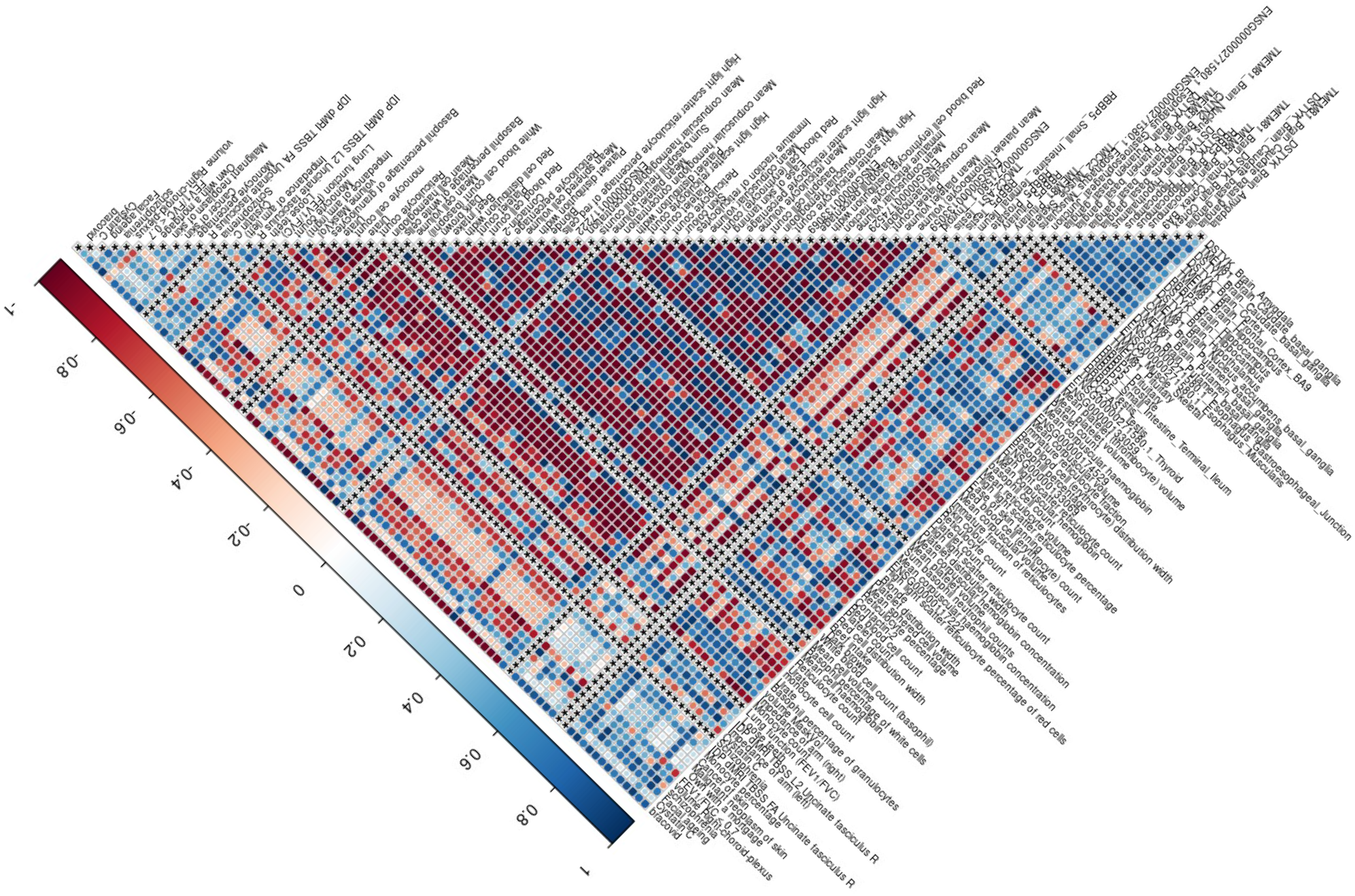
Pairwise colocalization results for all pairwise combinations of significant GTEx gene/tissue pairs, traits associated with rs11240388 in the phewas analysis and BRACOVID hospitalization results for the rs11240388 region. Red spectrum for colocalization results where H3 posterior probability > H4 posterior probability. Blue spectrum for colocalization results where H4 posterior probability > H3 posterior probability.

### Supplementary files

**Supplementary File 1**. Replication results from BRACOVID for HGI B1 analysis genome-wide significant loci.

**Supplementary File 2**. Replication results from BRACOVID for HGI B2 analysis genome-wide significant loci.

**Supplementary File 3**. Genome-wide significant hits from meta-analysis BRACOVID versus HGI B1 model.

**Supplementary File 4**. Genome-wide significant hits from meta-analysis BRACOVID versus HGI B2 model.

## References

1. Zhu N, Zhang D, Wang W, Li X, Yang B, Song J, et al. A Novel Coronavirus from Patients with Pneumonia in China, 2019. N Engl J Med. 2020;382(8):727–33.

2. COVID-19 Dashboard by the Center for Systems Science and Engineering (CSSE) at Johns Hopkins University (JHU) [Internet]. 2021. Available from: https://coronavirus.jhu.edu/map.html

3. Morens DM, Fauci AS. Emerging Pandemic Diseases: How We Got to COVID-19. Cell. 2020;183(3):837.

4. Casanova J-L, Abel L. Lethal Infectious Diseases as Inborn Errors of Immunity: Toward a Synthesis of the Germ and Genetic Theories. Annu Rev Pathol. 2021 Jan;16:23–50.

5. Casanova JL, Abel L. The human genetic determinism of life-threatening infectious diseases: genetic heterogeneity and physiological homogeneity? Hum Genet [Internet]. 2020;139(6–7):681–94. Available from: https://doi.org/10.1007/s00439-020-02184-w

6. Genomewide Association Study of Severe Covid-19 with Respiratory Failure. N Engl J Med. 2020;383(16):1522–34.

7. Pairo-Castineira E, Clohisey S, Klaric L, Bretherick AD, Rawlik K, Pasko D, et al. Genetic mechanisms of critical illness in COVID-19. Nature. 2021;591(7848):92–8.

8. Ganna A, Unit TG, General M. The COVID-19 Host Genetics Initiative, a global initiative to elucidate the role of host genetic factors in susceptibility and severity of the SARS-CoV-2 virus pandemic. Eur J Hum Genet. 2020;28(6):715–8.

9. Weinstein M, Skinner J. Covid-19 — Implications for the Health Care System David. N Engl J Med. 2011;362(5):567–71.

10. Allain-Dupré D, Chatry I, Michalun V, Moisio A. The territorial impact of COVID-19_J: managing the crisis across levels of government. OECD Tackling Coronavirus. 2020. p. 2–44.

11. Altshuler DL, Durbin RM, Abecasis GR, Bentley DR, Chakravarti A, Clark AG, et al. A map of human genome variation from population-scale sequencing. Nature. 2010;467(7319):1061–73.

12. Preparation S, No DP. Axiom ™ 2.0 gDNA Sample Preparation [Internet]. p. 2–5. Available from: https://www.thermofisher.com/us/en/home/technical-resources/contact-us.html?supportType=TS

13. Boughton AP, Welch RP, Flickinger M, VandeHaar P, Taliun D, Abecasis GR, et al. LocusZoom.js: Interactive and embeddable visualization of genetic association study results. Bioinformatics. 2021 Mar;

14. Barrett JC, Fry B, Maller J, Daly MJ. Haploview: analysis and visualization of LD and haplotype maps. Bioinformatics. 2005 Jan;21(2):263–5.

